# Single cell RNA-seq discovery of blood biomarkers predicting treatment outcome in severe asthma patients

**DOI:** 10.1101/2025.04.16.25325934

**Authors:** Lorena Rodrigues Sabino, Hui Ying Tan, Gabrielle Dziura, Isabella Mackay, Carlos Riveros, Peter AB Wark, Gerard E Kaiko

## Abstract

Biologic monoclonal antibody therapies for severe asthma target the Type 2 endotype through blockade of the IgE, IL-5/eosinophil, or IL-4/13 pathways, which represents at least two-thirds of patients, and have led to significant clinical benefits in severe asthma management. However, studies show that 10-20% of patients may be non-responders and require a change in therapy. There is also the emerging concept that a significant percentage of patients may enter ‘clinical remission’, with a very high level of disease control and virtually symptom-free. These clinical scenarios and heterogeneity increase the need to develop blood-based biomarkers that can predict outcome. Identifying markers of clinical remission may also have potential for expanding access to other severe asthma patients not currently identified through serum IgE, blood eosinophils, or FeNO. In this study, blood was taken prior to therapy from severe asthma patients (n=31) with a Type 2 endotype, high serum IgE, atopy, and blood eosinophilia who qualified for both Omalizumab (anti-IgE) and Mepolizumab (anti-IL-5) and were randomised to receive either treatment. White blood cells underwent single cell RNA-sequencing and patients were assessed for clinical outcomes over a 6-month period. Non-response to either Omalizumab or Mepolizumab was predicted by a gene signature expressed in antiviral plasmacytoid dendritic cells. Clinical remission was predicted by a common gene signature in rarer CD34+ blood progenitors and circulating MAIT cells with a ROC Curve AUC of 0.91 and 0.88, respectively. This discovery study identifies novel blood biomarkers that predict clinical outcome to multiple biologic therapies in severe asthma.

## Introduction

Severe asthma represents a significant global health burden, affecting millions of individuals worldwide and imposing substantial economic and healthcare costs [1]. While the majority of asthma patients achieve adequate control with conventional therapies, the severe asthma subset remains poorly controlled [2, 3]. Severe asthma poses a considerable challenge due to its complex and heterogeneous pathophysiology. Over the past few decades, significant advances have been made in understanding the underlying mechanisms driving severe asthma. Severe asthma is characterized by persistent symptoms, frequent exacerbations, significant airflow limitation, and poor quality of life despite adherence to high-dose inhaled corticosteroids (ICS) and long-acting beta-agonists (LABA) [4]. The majority of people with severe asthma, display, persitent type 2 inflammation, characterised by eosinophilic airway inflammation, and IgE-mediated responses that are relatively refractory to treatment with inhaled corticosteroids [2, 3]. These underlying mechanisms serve as therapeutic targets for biologic agents aimed at modulating specific components of the immune response, including Omalizumab (IgE), Mepolizumab and Reslizumab (IL-5), Benralizumab (IL-5 receptor), Dupilumab (Common IL-4/IL-13 Receptor), and Tezepelumab (Thymic stromal lymphopoietin) [5].

Mepolizumab, a monoclonal antibody targeting IL-5, and Omalizumab, a monoclonal antibody against IgE, are highly effective therapeutic options for severe asthma with the type 2 inflammation endotype [2]. Mepolizumab acts by inhibiting the biological activity of IL-5, thereby reducing eosinophilic inflammation, and is targeted at patients with blood or sputum eosinophilia [5, 6]. Omalizumab exerts its therapeutic effects by binding to free IgE and preventing its interaction with high-affinity IgE receptors on mast cells and basophils, and is thus best targeted at patients with high serum IgE [5, 6]. Clinical trials evaluating Mepolizumab and Omalizumab have demonstrated significant reductions in asthma exacerbations, oral corticosteroid (OCS) use, and improvements in asthma control and symptoms [5, 7, 8].

Despite the efficacy of Mepolizumab and Omalizumab in specific asthma phenotypes, not all patients respond to these therapies. Response rates vary depending on multiple factors, including those with features of uncontrolled Type 2 inflammation appear most responsive, while those with more comorbid chronic disease and who are obese are less likely to be responsive [9]. There is a lack of ability to accurately predict a patient’s non-response when they have already qualified to receive a biologic therapy [10, 11]. Moreover, access to biologic therapies remains restricted by their high cost and stringent eligibility criteria. Initiation of therapy requires careful clinical consideration of asthma severity, phenotype, exacerbation history, levels of blood eosinophil counts or serum IgE, and comorbidities. Furthermore, there is an important emerging post-biologic treatment paradigm of clinical remission in severe asthma [11, 12]. Patients that enter clinical remission (rates vary between 20-30%) are able to have their disease successfully controlled with minimal or no asthma symptoms, no exacerbations, and no OCS use after treatment with a biologic therapy plus maintenance ICS [11, 12]. Those in clinical remission can also substantially reduce their use of inhaled corticosteroids [13]. The ability to predict clinical remission with the same biomarker across multiple biologic therapies would provide a powerful clinical tool to potentially enable more patients at an earlier stage of severe asthma management to receive a biologic therapy. Therefore, this highlights the need for better patient stratification and precision treatment approaches using non-invasive biomarkers to predict clinical outcomes.

The advent of biologic monoclonal antibody therapies represented a paradigm shift in the management of severe asthma, offering targeted treatment options for patients with uncontrolled disease. However, the next step is optimising the use of these therapies, as well as potentially expanding this in a targeted manner to patients earlier on in the course of their disease potentially through the use of precise biomarkers. In this study we conducted a discovery analysis to identify peripheral blood cell biomarkers in severe asthma that predicted clinical outcomes after 6 months of biologic therapy. Blood samples from patients who were eligible to receive either Mepolizumab or Omalizumab (n=31, from a sub-study of a larger parallel arm, open label, randomized controlled clinical trial; ACTRN12618000850279) were taken prior to therapy initiation and single cell RNA-sequencing (scRNAseq) was conducted on white blood cell (WBC) fractions. We used multiple single cell data analysis pipelines combined with a post-hoc analysis of clinical trial data. Biomarkers were identified that predicted clinical outcomes (clinical remission, non-response, ACQ5, OCS use, and frequency of exacerbations) across both biologic therapies.

## Methods

### Patient selection and study population

This study was approved by the Hunter New England Local Health District Human Research Ethics Committee (2019/ETH01231) and written informed consent was obtained from each study subject. Inclusion criteria reflected Australian prescribing requirements and as previously described [14], Briefly these were; age ≥12 years; severe asthma despite high dose inhaled corticosteroid/long acting beta agonist and treated by a respiratory physician or immunologist. Evidence of poor asthma control in the past 12 months defined as: an acute exacerbation requiring treatment with OCS for at least 3 days, or requiring regular OCS daily for at least 6 weeks, with a cumulative dose of OCS of at least 500 mg prednisolone, and poor asthma symptom control, Asthma Control Questionnaire (ACQ-5) score of ≥2.0, as assessed in the month prior to the initial study visit,. Evidence of a dual allergic/eosinophilic phenotype defined as: total serum IgE >30 IU/mL, evidence of atopy documented by skin prick testing or positive radioimmunoassay to airborne allergens and blood eosinophil count ≥300 cells per microlitre in the 12 months prior.

### Randomisation

Randomisation was performed using the CReDITSS Online Randomisation Engine (CORE), a web-based randomisation and data entry system deployed as a standalone application for individual research projects. Permuted block randomisation with block sizes of 4 or 6, stratified by baseline eosinophil count (using a median split), was used. Eligible participants who provided informed consent were randomised (1:1) using the CORE system to receive either open-label Mepolizumab or Omalizumab for 6 months.

### Treatments

Mepolizumab and Omalizumab were prescribed with the dosage determined by the treating physician according to standard clinical care guidelines.

### Clinical outcomes

Baseline data, lung function tests and blood samples were collected by the local site at the time of randomisation and before administration of the biological agent. All participants were contacted monthly by phone to assess changes in clinical condition using the ACQ5 for symptom control [15, 16], record exacerbations (use of prednisone for more than 3 consecutive days), and monitor unplanned hospital visits for asthma exacerbations, or adverse events. At the end of the 6 month treatment trial, clinical assessments and blood samples were collected to determine treatment success or failure. Treatment success was determined by reduction in ACQ5 by at least 0.5 [15, 16] or reduction of total use OCS (prednisone) by at least 15%. Treatment was determined as failure or ‘non-response’ if there were no reduction in ACQ5 by at least 0.5, failure to reduce total use or maintenance OCS by 15%, an increase in exacerbations requiring hospitalisation, intolerance to the agent or significant adverse effects. Clinical remission was defined based on criteria published by Thomas et al. [12], with modifications for lung function criteria due to significant airflow obstruction in the cohort. Remission was defined as: ACQ5 score of less or equal to 1; no recorded exacerbations (use of prednisone for more than 3 days), no maintenance use of OCS, no hospitalisation for exacerbations, FEV1 stable or unchanged (had not fallen below 3% of the baseline FEV1). One patient in this study underwent treatment for Omalizumab and experienced a severe adverse event on first dose, and treatment was immediately ceased. The patient underwent a 1 month wash out period and then blood was re-sampled immediately prior to initiating Mepolizumab.

### Whole blood preparation for scRNA-seq

Human blood specimens were collected immediately prior to therapy initiation in EDTA coated tubes and processed for scRNAseq within 2 hours. Blood was kept at room temperature. A total of 4 mL of blood was obtained for whole blood preparation for single-cell RNA sequencing (scRNA-seq). Peripheral blood mononuclear cells (PBMCs) and granulocytes were separated using Mono-poly Resolving Medium (M-RPM. MPBio, 1698049) and Lymphoprep (Stem Cell Technologies, 07801) according to the manufacturer’s protocol. Briefly, 3.5 mL of blood from the stock tube was carefully layered on top of 3 mL of M-RPM in a 15 mL tube. This was then centrifuged at 300 x g for 30 minutes in a swinging bucket rotor at room temperature. Using a transfer pipette, the PBMC and granulocyte layers were carefully collected into a sterile 15 mL tube. The collected cells were then diluted in an equal volume of PBS with 2% FBS. This diluted sample was carefully layered on top of the Lymphoprep and centrifuged at 800 x g for 20 minutes at room temperature with the brake off. The upper plasma layer was discarded. The PBMC and granulocyte layers were collected and combined in a 2:1 ratio, washed twice with washing medium (High glucose DMEM, 10% FCS, 1% P/S, 1% L-glutamine) and centrifuged at 150 x g for 10 minutes at room temperature with the brake off to remove platelets. The supernatant was discarded, and the cells were resuspended in 5 mL washing medium for cell counting. One million of the resulting white blood cells from this combined cell population were collected and centrifuged at 300 x g for 5 minutes, then resuspended in 500 µL RBC lysis buffer for 4 minutes to remove any remaining RBC contaminants. After 4 minutes, 14 mL of washing medium was added, and the mixture was centrifuged at 300 x g for 10 minutes. The cell pellet was then resuspended in 700 µL washing medium and adjusted to a concentration of 1 million cells/mL. A 40 µL aliquot of the cell suspension was transferred to a 1.5 mL microcentrifuge tube, to which 1 µL of RNase inhibitor was added and proceeded to 10X Genomics 3’ NextGEM protocol and library preparation.

### scRNAseq on blood cells

Cells were processed with the 10XGenomics Chromium NextGEM v3.1 mRNA 3’ protocol as per manufacturer’s instructions. Briefly, single cells were partitioned into gel beads in emulsion with cell lysis and reverse transcription of RNA introduces cell barcoding. This step is followed by PCR amplification, clean-up, and double-sided size selection with SPRIselect reagent (Beckman Coulter), and adaptor ligation and sample indexing by PCR. We aimed to recover a maximum of 10,000 cells per patient sample. Samples were mixed at an equimolar ratio after indexing and sequenced with a Novaseq 6000 S4 200 cycle flow cell followed by de-multiplexing.

### scRNAseq bioinformatic analysis pipelines

Following demultiplexing using the indexes from libraries to inidvdiual patient samples, the reads were aligned to different references using two different pipelines. CellRanger, which is a standard 10X Genomics designed analysis pipeline that aligns using STAR and maps to the whole genome [17]. AlevinFry is a new method, which maps against a reference that indexes both the alternative spliced transcriptome and the set of (collapsed) intron sequences with a spliced + intronic reference [18]. This method allows more read mapping to the spliced transcriptome. The distribution of droplet (cell) barcodes ranked by total unique molecule identifier (UMI) barcodes is used by both CellRanger and AlevinFry to determine a threshold that distinguishes proper cell barcodes from empty droplets, albeit each pipeline uses a different strategy. The number of barcodes included is based on a similar algorithm for AlevinFry,

Cellranger and emptyDrops, but the higher mapping efficiency for AlevinFry makes for extended limits [19]. In both cases estimating the non-empty barcodes, provides an estimate of the ambient RNA from all barcodes with low UMI counts. Automatic CellRanger threshold produced cell counts below the target number of cells, but closer inspection of the UMI – barcode rank plots and comparison to AlevinFry results revealed that Cellranger parameters were too stringent, and a final mapping was run forcing Cellranger target number of cells. Both alignment pipelines were used as implemented in the curated nf-core “scrnaseq” v2.3.2 nextflow workflow [20]. The single cell count data stored in HDF5 (Cellranger) or Seurat (Alevin-Fry) file formats for later analysis in R v4.3.3. Extensive QC with quantification of mitochondrial genes, computation of per-barcode statistics: UMI counts, genes detected, proportion of counts in mitochondrial genes, rank was performed. Based on the analysis, cells (barcodes) with less than 50 total UMI counts and cells with 20% or higher of total UMI counts in mitochondrial genes were excluded from posterior analysis, but barcodes with less than 50 UMI counts were used to estimate ambient RNA contribution for Monte-Carlo refinement of empty cell likelihood. Additional quality control checks included doublet cell barcode detection, low UMI count outlier detection, low number of genes per barcode detected and exclusion of genes detected in 10 cells or less. Final numbers for the 31 samples were: cells AlevinFry: (mean 9941.5, [9013–10629]), CellRanger: (mean: 8735.6 [4407–10002]), genes: AlevinFry: (mean: 36925.9 [31257–43888]), CellRanger: (mean: 17121.9 [18392–19717]). Single cell data for each alignment method for all samples were then integrated in a single dataset for consistent cell type identification and clustering analysis using the R packages scran, scater and batchelor and Secuer [21, 22]. Clustering results in the integrated dataset were compared with individual sample clustering, and consistency was also checked with alternate clustering methods (walktrap, Leiden). Cell type identification was performed on a cell-by-cell basis using the SingleR package with a combined cell type reference database made of the ENCODE, Monaco and Human Protein Cell Atlas. The combined cell type reference provided cell labelling more consistent with the identified clusters than any single individual reference.

The cell type labels associated to each barcode from this analysis were then transferred back to the pre-integration individual sample data to obtain consistently annotated cell-type-labelled single cell data used in per sample differential gene expression analysis between cell types. The annotated single cell datasets were further transformed into cell-type-annotated pseudo-bulk transcriptomic data by aggregating gene counts across cells for each cell type present in the sample. Pseudo-bulk values of cell types with less than 20 cells in the sample were labelled as NA. The pseudo-bulk datasets were used in the biomarker univariate and multivariate analysis as described below.

### Cell type and sample differential gene expression and statistical models

Two different differential expression analysis were performed on the cell-type annotated data: i) a per-sample single cell differential expression between cell types, and ii) a differential expression analysis on pseudo-bulk cell type data with clinical outcomes as response variables, with and without age and sex as covariates. The per-sample analysis uses (normalised) log counts with cell type labels from previous cell type identification in the integrated data set. The differential expression uses two methods: a Wilcoxon cell type pairwise comparison combined across all cell type pairs to obtain the most significantly upregulated genes in a cell type (as implemented in the findMarkers and combineMarkers functions in the R package scran v1.30.2), and a marker scoring method that summarises effect sizes by combining simple statistics on standardised log-fold changes and AUC for each gene across pairwise comparisons involving a cell type of interest (also implemented in the scran package function scoreMarkers).

The clinical outcome differential expression analysis uses the per-sample pseudo-bulk gene expression data generated as explained before. From the total 22 (AlevinFry) or 21 (CellRanger) identified cell types, a median of 17 (AF) or 16 (CR) types were present in any one sample. The differential expression is performed with the edgeR and scran R packages [23], and models were built with treatment and sample as blocking factors for all clinical outcomes of interest; change in OCS, exacerbations, ppFEV1, Responders vs. Non-responders, Clinical Remission vs others, and adverse events. Age and sex were tested for inclusion as adjustment covariates. The top 500 cell type – gene most performant results of the clinical differential expression analysis were further used for biomarker discovery. Both univariate and multivariate models were constructed on each clinical outcome variable, and candidate cell type – gene results ranked by their individual univariate ROC AUC performance on a 3:1 training: test split of sample’s data. Univariate ROC AUC for logistic (categorical outcomes) or generalised regression (continuous / ordinal outcomes) was used to estimate individual cell-type – gene predictive power. Finally, to estimate performance of biomarkers related to clinical outcomes, we trained random forests and elastic net multivariate logistic or regression predictor models, assessing their performance by multiple metrics [24]. The elastic net parameters were set as mixture = 0.99 (almost a pure LASSO model) and penalty = 0.001 through a few initial runs and used unchanged throughout. The random forest model parameters were 1000 trees with a minimum node size of 5. The models were trained and tested on a 2:1 split of the data. Metrics used for performance evaluation were: R squared, root mean squared error, Huber loss, ratio of performance to deviation, ratio of performance to inter-quartile for regression models / continuous outcomes, and ROC AUC, mean precision, precision – recall AUC, gain curve AUC, Brier class, accuracy, precision, recall, Kappa, F1 measure and Matthews correlation coefficient. All modelling was performed in R v4.3.3 using packages tidymodels v1.2.0, parsnip v1.2.1, glmnet v4.1.8, randomForest v4.7-1.1 [25, 26].

## Results

### Improvement in clinical outcomes following treatment with either Omalizumab or Mepolizumab

This scRNAseq discovery clinical data analysis represents a sub-study on a subset of patients from a larger Phase 4, parallel arm, open label randomized controlled clinical trial Choosebetweenamab (ACTRN12618000850279). A discovery analysis was conducted on WBCs from a severe asthma patient cohort (n=31 samples) using patient blood taken prior to initiation of treatment with either Omalizumab or Mepolizumab. This sub-study was an exploratory investigation for biomarker discovery using scRNAseq and is not designed to detect head-to-head clinical outcome differences in the treatment arms, which is the purpose of the larger Phase 4 trial. However, in order to demonstrate the range of responses in this sub-study we conducted post-hoc analysis of any differences in clinical outcomes between treatment arms or as a joint analysis of both Mepolizumab and Omalizumab. Table 1 shows the patient demographic and baseline clinical characteristics in this study. Groups were age– and sex-matched. For the purposes of this discovery study, we assessed changes after 6 months in; ACQ5, OCS use, frequency of exacerbations, and lung function (ppFEV1). We also assessed these changes as a % of the patient’s baseline measure. In this sub-study we found no real significant differences in clinical outcomes between Mepolizumab, Omalizumab (Figure 1A-H and Table 1). As expected from both Phase 3 clinical trial data and post-approval studies the majority of patients (∼85%) experienced improved clinical outcomes (Response) for either therapy and continued to be maintained on these therapies (Figure 1I). We measured the number of adverse events (drug or non-drug related). Adverse events that could be definitively related to drug were too low in frequency for analysis. Adverse events were almost exclusively minor and occurred in similar percentages of patients for either therapy (Figure 1J). Clinical remission was defined asACQ5 of <1, no exacerbations over 6 months, no use of OCS, and no decline in lung function. Clinical remission was detected in 29% of severe asthma patients in the overall joint analysis (22% Mepolizumab and 38% Omalizumab) (Figure 1H).

**Figure 1.**
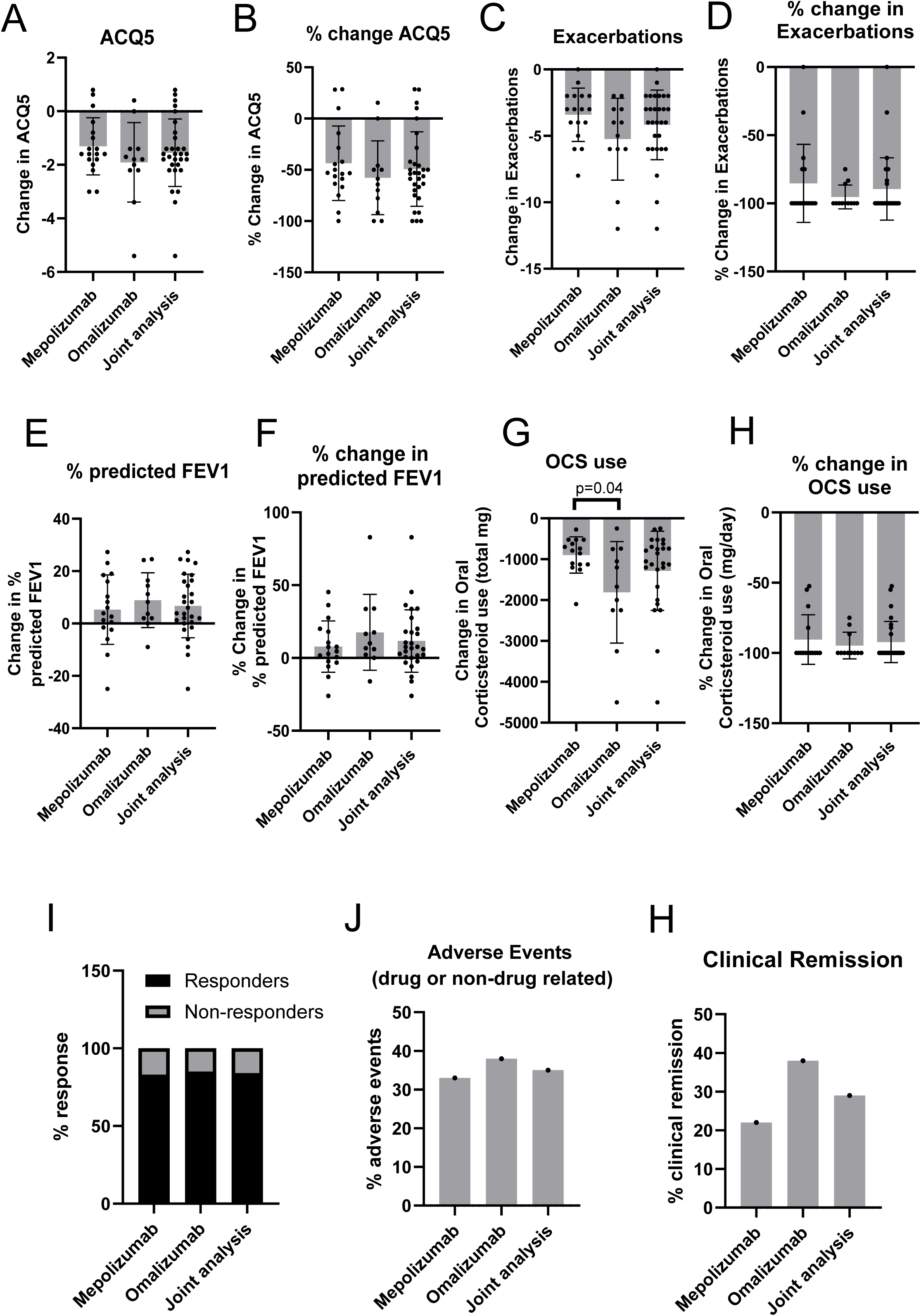
Clinical outcomes for Mepolizumab, Omalizumab, and joint analysis. Severe asthma patients eligible for either Mepolizumab or Omalizumab therapy were randomised into either treatment and followed for 6 months with collection of clinical outcomes at baseline prior to initiation of therapy, and after 6 months of receiving either Mepolizumab (n=18), Omalizumab (n=13) or joint analysis of both (n=31). Asthma control questionnaire (ACQ5) was used to measure symptoms and expressed as an absolute change (A), and % change over baseline (B). Frequency of exacerbation up to 1 year prior to therapy initiation and during the 6 months of treatment were measured as absolute change (C), or % change over baseline (D). Lung function was assessed by percentage predicted FEV1 and quantified as absolute change (E), or % change over baseline (F). Oral corticosteroid (OCS) use in average mg/day was determined prior to therapy initiation and during the 6 months of treatment and expressed as absolute change (G), or % change over baseline (H). (I) The percentage of non-responders compared to responders (clinical response or clinical remission) was determined as a categorical outcome. (J) Percentage of patients experiencing adverse events on different treatments including related or unrelated to the drug. *p<0.05 by an unpaired t test.

**Table 1:** Baseline patient demographics and clinical characteristics.

|  | **Mepolizumab** | **Omalizumab** | **Joint Analysis** | **p value**<br>**Mepolizumab**<br>**Versus**<br>**Omalizumab** |
| --- | --- | --- | --- | --- |
| **Sample number** | 18 | 13 | 31 |  |
| **% female** | 72 | 62 | 68 |  |
| **Age (Mean ± SD)** | 51 ± 22 | 54 ± 17 | 52 ± 20 | 0.76 |
| **ACQ5 baseline**<br>**(Mean ± SD)** | 2.9 ± 0.6 | 3.1 ± 1.1 | 3.0 ± 0.8 | 0.42 |
| **OCS use baseline**<br>**(prednisone or**<br>**equivalent)**<br>**(Mean ± SD)** | 980 ± 406 | 1849 ± 1196 | 1266 ± 942 | **\* 0.03** |
| **% predicted**<br>**FEV1 baseline**<br>**(Mean ± SD)** | 84.3 ± 14.6 | 68.7 ± 24.4 | 77.6 ± 20.7 | **# 0.04** |
| **Exacerbations in**<br>**the prior 12**<br>**months (Mean ±**<br>**SD)** | 3.7 ± 1.9 | 5.5 ± 2.9 | 4.5 ± 2.5 | 0.08 |
| **Blood**<br>**eosinophils**<br>**baseline**<br>**million/ml**<br>**(Mean ± SD)** | 0.43 ± 0.24 | 0.64 ± 0.29 | 0.52 ± 0.28 | 0.05 |
| **Serum IgE**<br>**baseline U/ml**<br>**(Mean ± SD)** | 799 ± 1058 | 831 ± 1257 | 812 ± 1125 | 0.70 |
| **FeNO baseline**<br>**ppB (Mean±SD)** | 40.3 ± 28.8 | 22.6 ± 12.8 | 32.6 ± 24.6 | 0.10 |

\* p<0.05 by Mann-Whitney t test for non-parametric data

### Baseline clinical measurements before initiation of biologic therapy are poor predictors of clinical remission

We first determined in our clinical cohort whether blood eosinophils, serum IgE, and exhaled nitric oxide (FeNO) under joint analysis of all patients receiving either Mepolizumab or Omalizumab (n=31), predicted clinical remission. Using univariate analysis none of these biomarkers or clinical parameters accurately predicted response versus non-response or clinical remission within this overall joint analysis (receiver operating characteristic (ROC) curve analysis for all parameters, area under the curve (AUC) of <0.77 and none were statistically significant) (Figure 2A and Supplementary Table 1). To determine whether any of these clinical or biological parameters could predict the degree of response for individual clinical outcomes we conducted univariate linear regressions and assessed the r-squared values. For ACQ5 change over 6 months of therapy the only parameter that statistically correlated with this was the baseline ACQ5 score (Figure 2B). Similarly, change in OCS and change in exacerbations both had highly significant r^2^ value with baseline OCS use and baseline number of exacerbations (Figure 2B and Supplementary Table 1), respectively. There were no significant correlations between change in FEV1 and any baseline metrics. However, when the change in the clinical outcomes over 6 months was converted to the % of that individual patient’s baseline for the same outcome these statistically significant correlations with ACQ5, OCS, and exacerbations no longer held true (Figure 2C and Supplementary Table 1). This suggests these individual clinical outcome r^2^ correlations may have a mathematical basis rather than a biological basis. Only relatively weak correlations emerge when the change in clinical outcome data is converted to a % of baseline showing that baseline FEV1 and FVC correlate with the % change in predicted FEV1 (r^2^=0.26-0.28, p<0.01) (Figure 2C and Supplementary Table 1).

**Figure 2.**
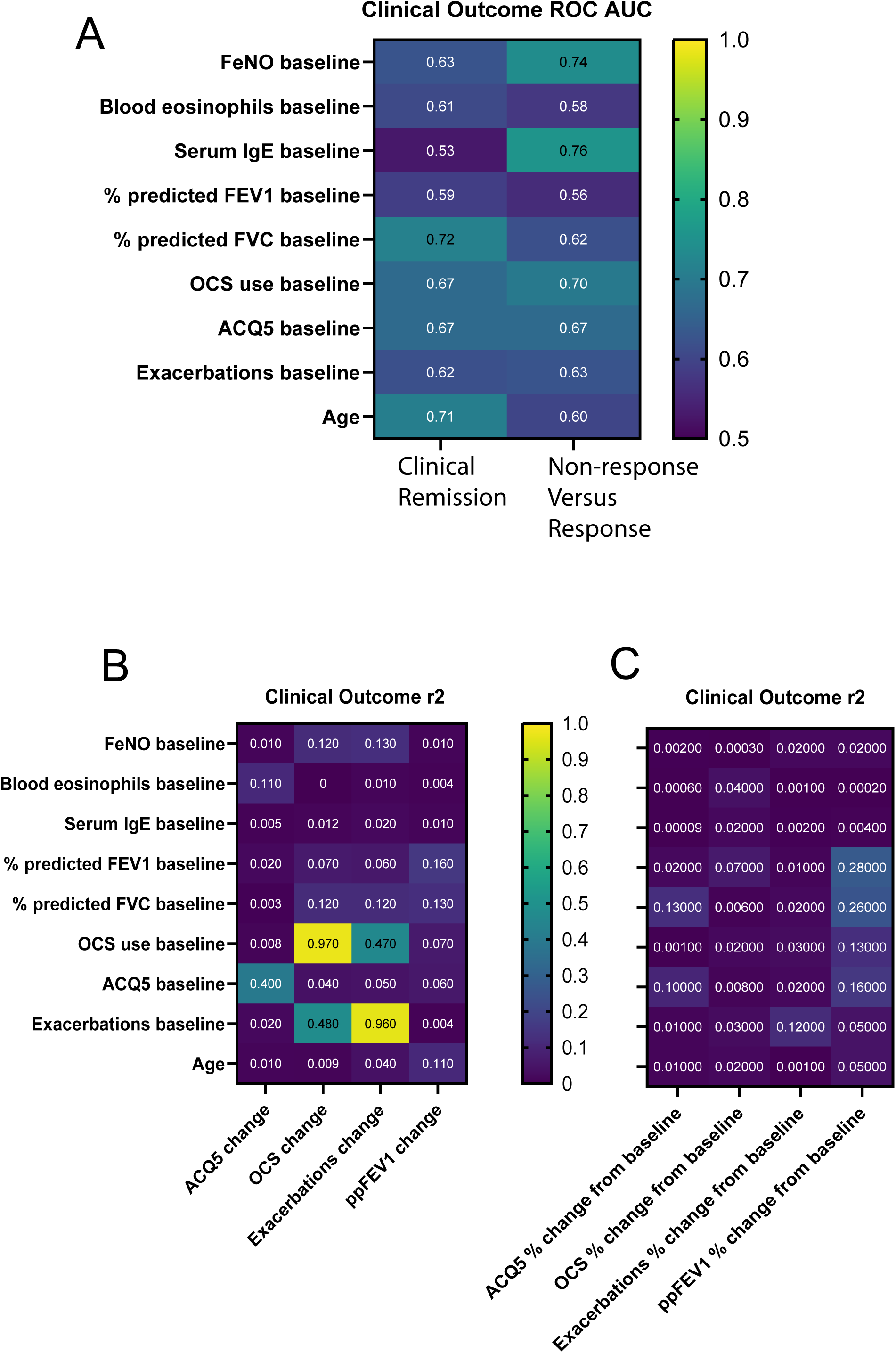
Association of clinical baseline characteristics with treatment outcomes after 6 months in the joint analysis for patients receiving either Mepolizumab or Omalizumab. (A) Receiver operating characteristic (ROC) curve analysis for ability of baseline clinical characteristics of severe asthma patients (n=31) to predict categorical clinical outcomes of clinical remission, or non-response to therapy. Box text indicates area under the curve (AUC) scores. Linear regression correlations of baseline characteristics with clinical outcomes expressed as either absolute value changes from baseline (B), or relative % change from baseline (C). Box text indicates r^2^ correlation coefficient.

### scRNAseq on white blood cells before initiation of biologic therapy with data analysis by two independent methods

Our data demonstrates that better biomarkers are needed to predict clinical remission. Given the relative homogeneity of patients before therapy initiation in a severe asthma population on similar medications who all meet the same clinical requirements for access to these biological therapies, we determined that a discovery technology with greater resolution would be required than traditional approaches such as bulk RNA-seq. On this basis we employed scRNAseq on white blood cells isolated from patients immediately prior to initiation of Mepolizumab or Omalizumab (Figure 3A). This technology is able to detect differences in gene expression (especially in rarer immune cell types) that would otherwise be masked by the averaging effect of bulk sequencing approaches. Both granulocytes and mononuclear cell fractions were isolated from patient blood and then combined. Each sample had approximately 10,000 cells sequenced by the 10X Genomics NextGem pipeline with 25,000 reads/cell, libraries were created, QC conducted, and then Illumina sequencing with Novaseq. We applied two independent bioinformatic pipelines to the analysis of the resulting scRNAseq data. The first pipeline used the CellRanger methodology that identifies cell types and gene expression based on matching sequenced barcodes to tagged 3’ transcript sequences, whereas the AlevinFry methodology is able to identify splice variants of gene transcripts not possible with CellRanger by using a different reference genome approach that incorporates mapping to intergenic regions. Pseudo-bulk integration of data was conducted with all ∼310,000 single cells to create a uniform sample-to-sample approach to clustering and cell type annotation across all 31 samples. This was visualised using standard TSNE and UMAP plots (Figure 3B-E). We were able to detect approximately 20 different cell types using these approaches with small differences in the detection of some very rare cell types between CellRanger compared to AlevinFry. Quantification of cell type numbers by each method identified significant differences, albeit relatively small magnitude differences, in conventional dendritic cells, B cells, plasmacytoid dendritic cells, δγ T cells, Mucosal Invariant T (MAIT) cells, and CD34+ multipotent progenitor cells (Figure 4A). In each case AlevinFry identified a high number of these cell types compared to CellRanger.

**Figure 3.**
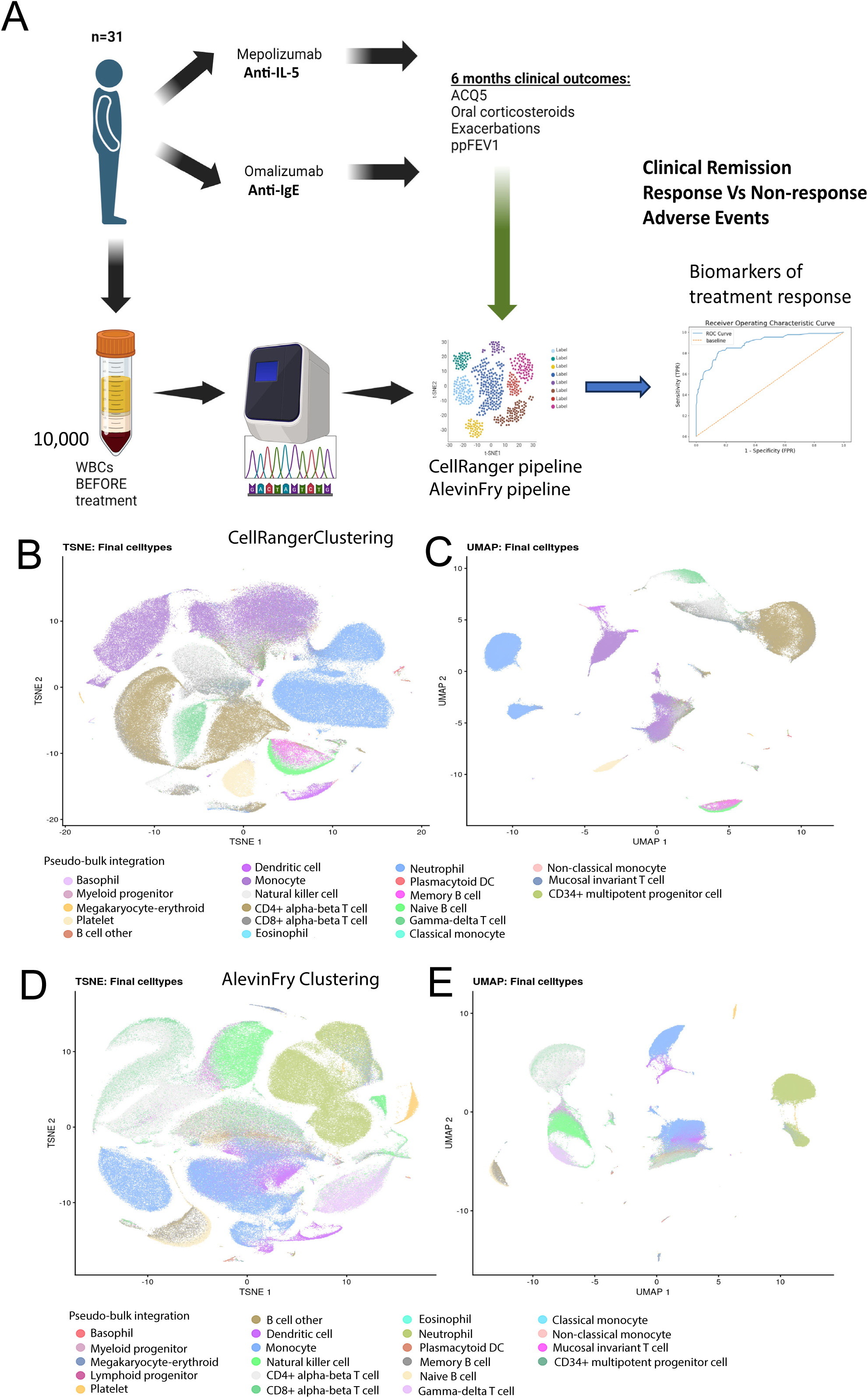
Single cell RNA-sequencing biomarker workflow employing two different analysis pipelines using white blood cells taken prior to initiation of therapy. (A) Severe asthma patients (n=31) were randomised to receive either Mepolizumab or Omalizumab and blood was taken prior to therapy initiation. Single cell RNA sequencing (scRNAseq) was conducted on ∼10,000 white blood cells per patient. Clinical data was collected across 6 months and this data was compared to scRNAseq signatures to identify biomarkers. (B) TSNE plot and (C) UMAP visualisation of pseudo bulk-integrated scRNAseq data from patient blood samples (n=31) used to identify clusters and cell type annotation with the CellRanger analysis pipeline. (D) TSNE plot and (E) UMAP visualisation of pseudo bulk-integrated scRNAseq data from patient blood samples (n=31) used to identify clusters and cell type annotation with the alternative AlevinFry analysis pipeline.

**Figure 4.**
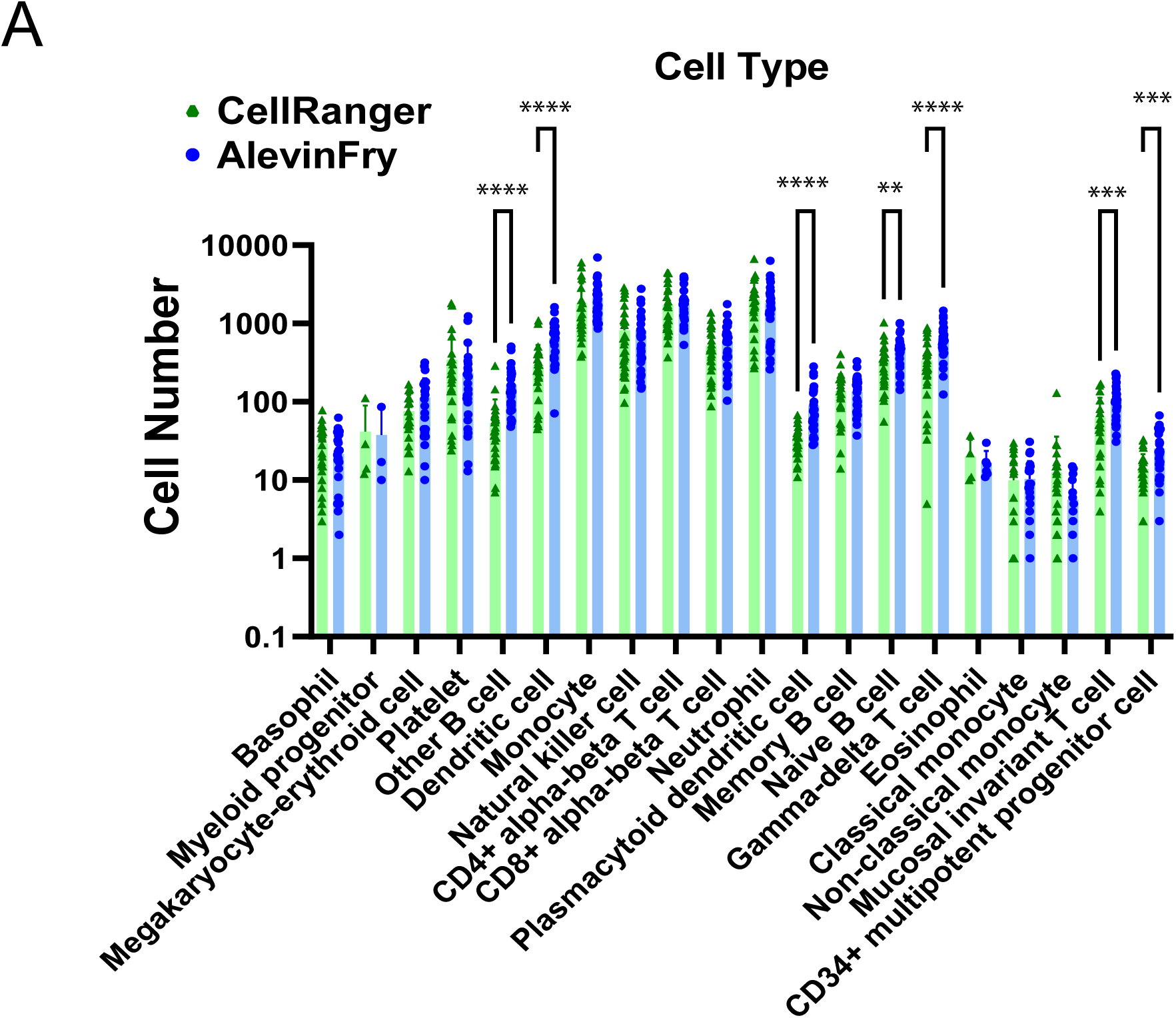
Blood cell type quantification using scRNAseq. (A) Cell type quantification from scRNAseq data of WBCs using two different pipeline with CellRanger and AlevinFry, **p<0.01, ***p<0.001, ****p<0.0001 with Mann-Whitney t tests and multiple comparisons correction using the Two-stage step-up (Benjamini, Krieger, and Yekutieli).

Typically, scRNAseq has difficulty detecting neutrophils due to their low RNA abundance and relatively high presence of RNAse. To overcome this our method was designed in several ways to allow the detection of neutrophil gene expression. This included blood collection in EDTA-coated tubes to prevent neutrophil activation, all steps performed at room temperature, an RNAse inhibitor added to WBCs, and an extra PCR cycle was added during library preparation for low abundance reads. This approach was highly successful (Figure 3 and 4), with an average of ∼2000 neutrophils detected per sample.

### scRNAseq quantification of blood immune cell type in severe asthma patients prior to treatment who later go on to clinical remission

We conducted joint analysis (n=31) on clinical remission (or response versus non-response) as there is greater clinical significance to identify blood-based biomarkers predicting outcome, in particular clinical remission, that are predictive for multiple biologic therapies. Furthermore, the joint analysis is necessary to provide sufficient sample numbers as some clinical outcomes have too low an occurrence to be analysed by individual therapy. As a global cell type analysis we compared all immune cell numbers in the patient blood just prior to initiation of biologic therapy with their 6-month clinical outcome of clinical remission or not. The data showed that there were no significant differences in total cell numbers by scRNAseq for any immune cell type in the blood of patients who would later go on to clinical remission (Figure 5A and B).

**Figure 5.**
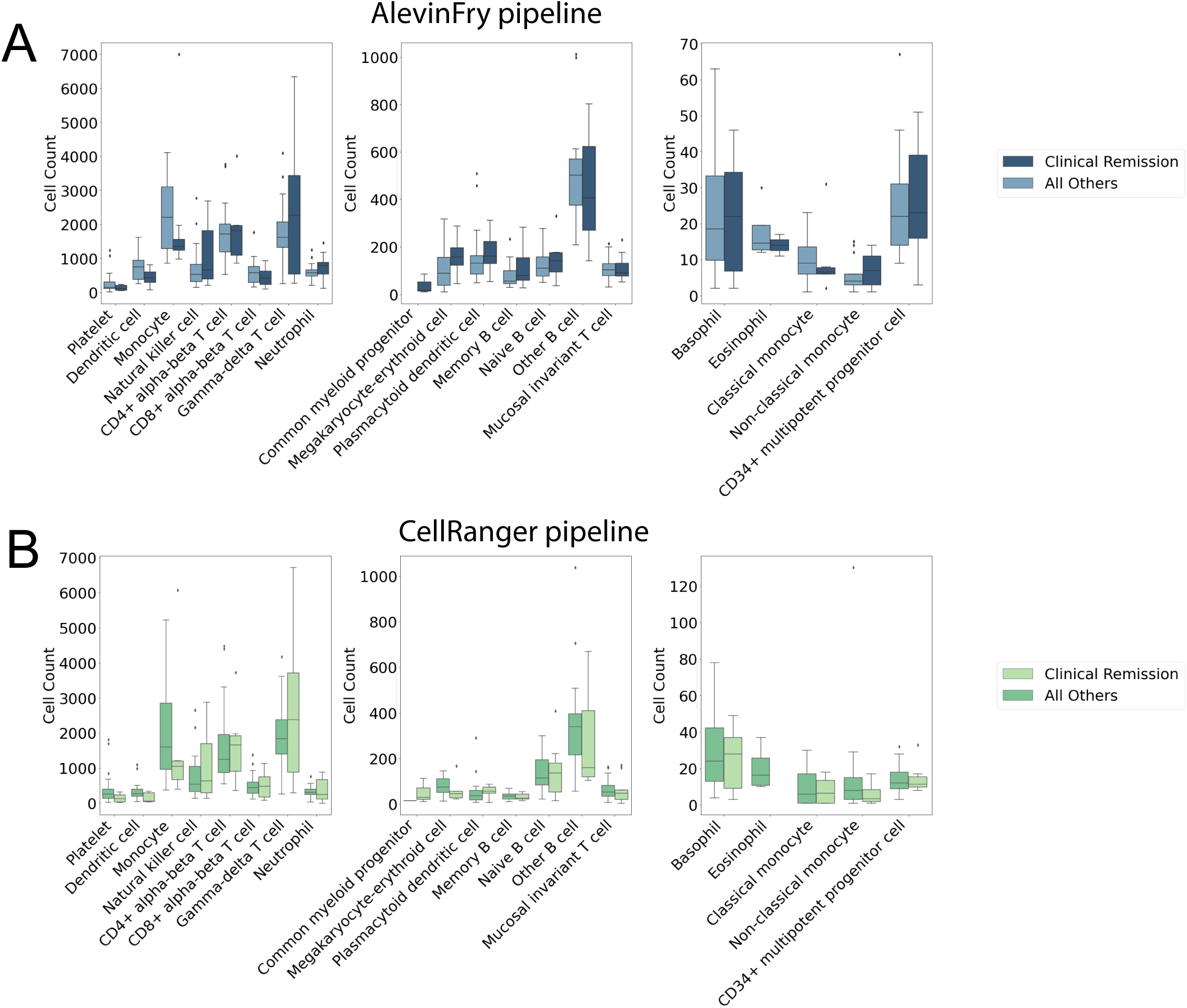
scRNAseq cell number quantification in blood prior to therapy initiation in patients who enter clinical remission or non-responders during the 6 month treatment phase. Cell type quantification from scRNAseq data of WBCs using two different pipeline with (A) AlevinFry and (B) CellRanger for clinical remission versus all others. All comparisons were non-significant using the Mann-Whitney t test and multiple comparisons correction using the Two-stage step-up (Benjamini, Krieger, and Yekutieli).

This analysis was repeated for both AlevinFry and CellRanger scRNAseq methodologies and the same results were observed in both cases indicating a high level of consistency in these findings between different pipelines (Figure 5A and B). No other statistically significant differences or correlations were observed in the total cell number for any immune cell types with any of the other clinical outcomes. These results suggest that total numbers of immune cell types, by themselves are relatively similar between all severe asthma patients eligible for biologic therapy prior to initiation and do not correlate with clinical outcomes.

### Gene activity in CD34+ progenitors and circulating MAIT cells is a highly accurate predictor of clinical outcomes, including clinical remission

Following identification and quantification of blood immune cell types in the severe asthma patient cohort (n=31) we investigated gene expression on an individual cell type basis. Using the cell types identified in Figure 3, we conducted differential gene expression in a pairwise fashion between clusters/cell types, and then compared this across all samples with regard to classification of clinical remission versus all other outcomes (response without clinical remission or non-response). From each differential expression result (outcome variable), we sorted genes in cell types by FDR and filtered for the top 500 gene-cell combinations. We then used a univariate model with ROC analysis to identify single cell type-gene expression predictors of clinical remission. Data was adjusted for age and sex differences between clinical outcome groups as well as analysed in a non-adjusted form. In order to identify conserved cell type-gene predictors with high confidence a ROC area under the curve (AUC) of ≥ 0.8 for predicting clinical remission was used and compared for commonality between four models including; sex/age-adjusted and non-adjusted data, as well as between CellRanger and AlevinFry scRNAseq methodologies. Using Venn diagrams these four comparisons for predictors of clinical remission resulted in 2 cell type-gene combinations that were highly predictive. These included CD34+ progenitor cells expressing *RPS26*, and blood circulating MAIT cells expressing *RSP26* (Figure 6A). Individual ROC curves show CD34+ progenitor cells-expressing *RPS26* to have an average accuracy across the four models with AUC of 0.91, and MAIT cells-expressing *RPS26*, to have an average accuracy of AUC 0.88 (across all 4 comparisons) (Figure 6B-C). We also conducted a univariate random forest approach using the top 500 ranked genes (with 1000 branches) and used the Mean Decrease in Gini Coefficient as the primary outcome metric. This analysis similarly identified CD34+ progenitor expressed *RPS26* as the top predictor of clinical remission (Figure 6D and Supplementary Table 2). We then used the top 100 ranked genes from each analysis component of the Venn diagram in Figure 6A (400 in total) and conducted over-representation pathway analysis for gene ontology (GO) terms to identify molecular pathways of interest linked to the clinical remission phenotype. This identified that the most highly enriched biological and molecular processes within the genes linked to clinical remission included apoptosis, cytokine-mediated signalling, as well as amino acid biosynthesis (Figure 6E). This suggests that there may be fundamental cellular activity differences in immune cells from severe asthma patients more likely to enter clinical remission from biologic therapy. *RPS26* is a ribosomal protein that is a general marker for cellular activity i.e. increased expression with increased cell proliferation and protein production. The data shows that average expression of this marker on a per cell basis is dramatically reduced in both CD34+ progenitors (4-fold down) and MAIT cells (3-fold down) of patients who go on to enter clinical remission (Figure 6F-G). Not only are these two cell type-gene combinations highly consistently predicted across methods, but they also have among the highest ROC AUCs for any individual method. These data show that lower *RPS26* gene expression in CD34+ progenitors or circulating MAIT cells is highly predictive of clinical remission in blood taken from patients prior to initiation of either Mepolizumab or Omalizumab.

**Figure 6.**
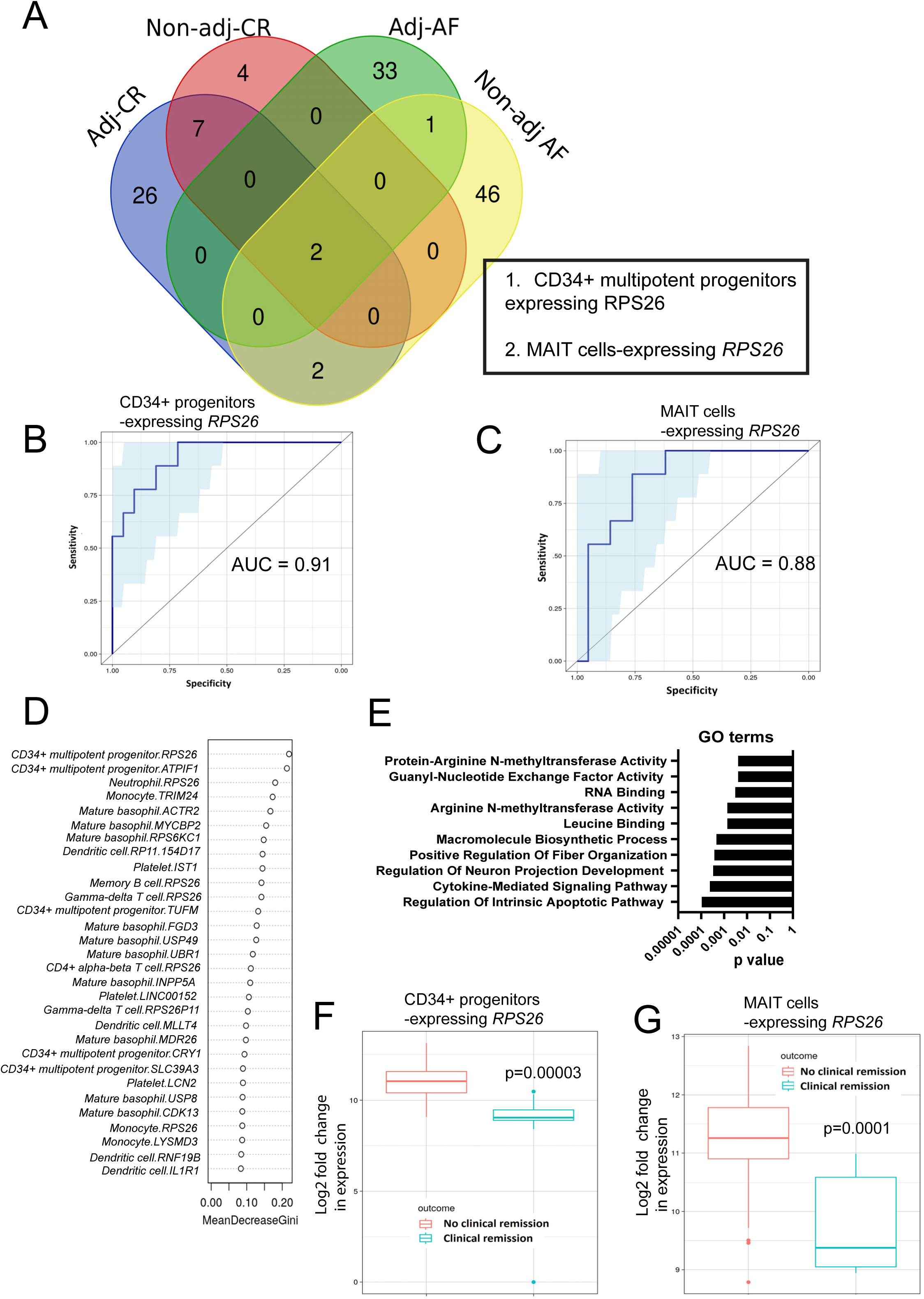
scRNAseq discovery of blood biomarkers predicting clinical remission for patients receiving either Mepolizumab or Omalizumab. (A) Venn diagram analysis for common biomarkers among 4 different comparisons of the cell type-gene features that have univariate ROC curve AUCs ≥ 0.8 for predicting clinical remission in patients receiving either Mepolizumab or Omalizumab (n=31). Biomarker sensitivity and specificity analysis by ROC curve for (B) CD34+ progenitors-expressing *RPS26* and (C) MAIT cells-expressing *RPS26*, with 90% confidence intervals (blue shading). (D) Random forest analysis of top biomarkers using the mean decrease in the Gini coefficient as the metric of each features contribution to the prediction of clinical remission from the model with the top marker in red box. (E) GO term analysis of the biological and molecular processes enriched in the genes that were most linked to clinical remission. Differential expression analysis as log2 fold change with p values indicated for the top and most consistent biomarkers predicting clinical remission (F) CD34+ progenitors-expressing *RPS26* and (G) MAIT cells-expressing *RPS26*. ROC; receiver operating characteristic (ROC), AUC; area under the curve. Adj-CR; age/sex adjusted data for Cell Ranger, Non-adj-CR; non-adjusted data for Cell Ranger, Adj-AF; age/sex adjusted data for Alevin Fry, Non-adj-AF; non-adjusted data for Alevin Fry, GO; gene ontology.

### An array of genes expressed in anti-viral plasmacytoid dendritic cells predict non-response to either biologic therapy

In our small discovery cohort ∼15% of severe asthma patients receiving either Mepolizumab or Omalizumab did not respond (Figure 1I). Previous large cohort studies have similarly shown this to vary between 10-20% [11, 27]. We investigated blood immune cell markers that may predict this lack of response. Again, we compared this across all patient samples with regard to categorisation of non-response versus all other outcomes (response without clinical remission or clinical remission). From the differential expression analysis, we identified the top 500 gene-cell combinations ranked by FDR. We then employed a univariate ROC model to identify cell type-gene expression predictors of non-response. In order to identify conserved cell type-gene predictors with high confidence a ROC AUC of ≥ 0.8 for predicting non-response was compared between the four models; adjusted and non-adjusted data for CellRanger and AlevinFry methodologies. Using Venn diagrams these four comparisons for predictors of non-response identified one cell type, the plasmacytoid dendritic cell (pDC), expressing many different genes as the dominant predictors of non-response. In particular, 5 genes (*ANXA2, CTSZ, RAB1B, DYNLL1,* and *ACP1*) were common between all four model comparisons (Figure 7A). However, many other pDC genes were found in this analysis and some possessed even higher ROC AUCs than the latter 5 genes including the pDC genes *DCAF5, IMMP2L,* and *FAF1* all with ROC AUCs of 1.0. For example, individual ROC curves showing pDCs-expressing either *ANXA2* or *DCAF5* have an accuracy by AUC of 0.89 and 1.0 in the AlevinFry (age/sex-adjusted analysis), respectively (Figure 7B-C). Given that total pDC numbers were not different between responders and non-responders, this data suggests that it is the transcriptional activity or activation status of pDCs that is altered in patients who will go on to be non-responders. We then conducted a univariate random forest approach using the top 500 ranked genes (with 1000 branches) and used the Mean Decrease in Gini Coefficient as the primary outcome metric. This analysis similarly identified almost exclusively pDC genes as the top predictors of non-response including validating those already identified by univariate ROC scores; *DCAF5, IMMP2L, and FAF1* (Figure 7D). In fact, greater than 99% of cell-gene combinations in the top 100 ranked by ROC AUC by all four methods of analysis (Figure 7A) were genes expressed in pDCs. We then conducted GO term pathway analysis, which showed that NADH mitochondrial metabolic processes, as well as proteasomal and ubiquitination pathways were enriched in the pDC genes linked to non-response (Figure 7E). Furthermore, we found that almost all pDC genes with a ROC AUC ≥ 0.8 for predicting non-response are downregulated in non-responder patients, suggesting there is a loss of pDC gene activity in non-responders (Figure 7F). For example, *DCAF5* is downregulated ∼3-fold in pDCs of patients who go on to non-response (Figure 7G).

**Figure 7.**
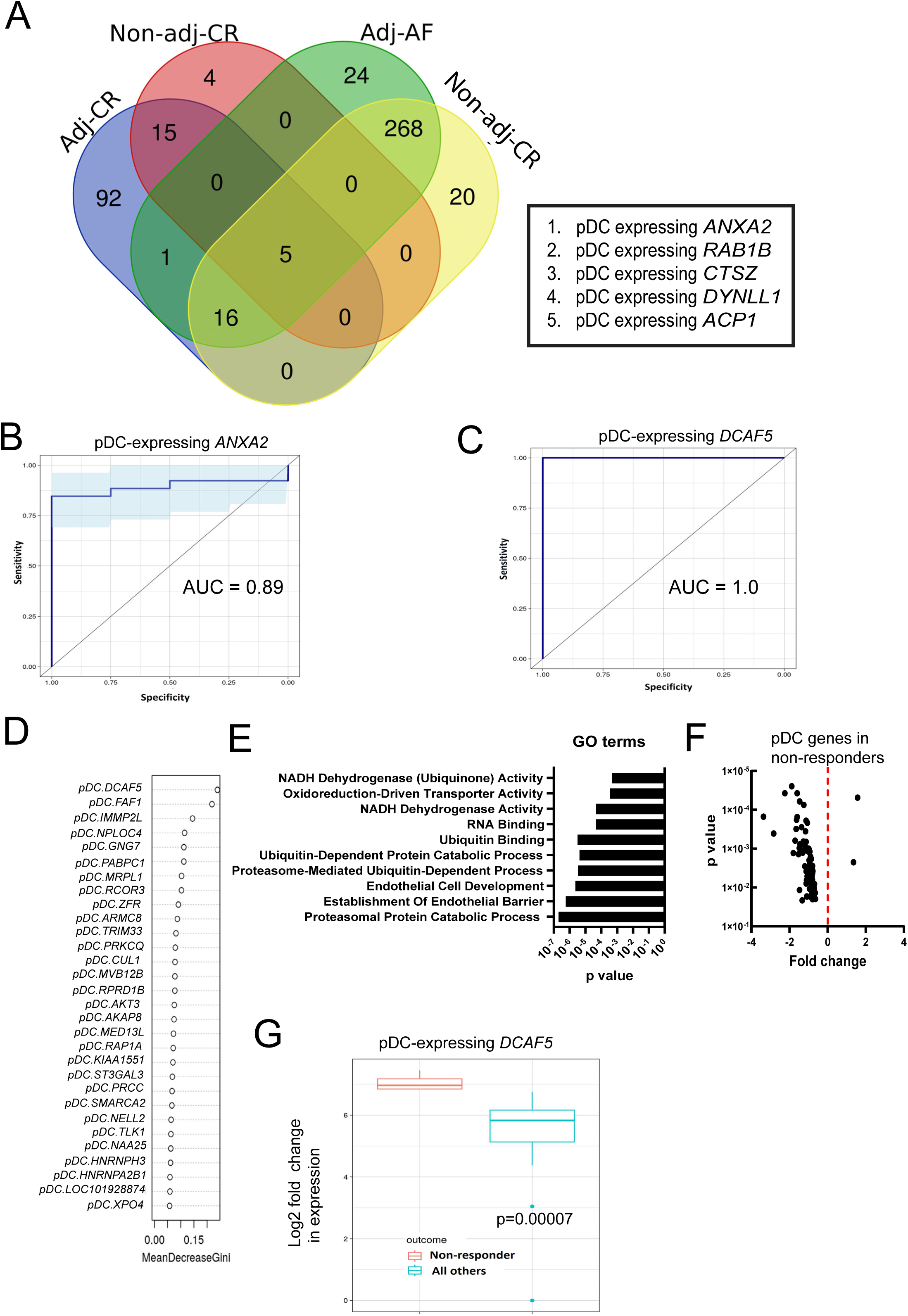
scRNAseq discovery of blood biomarkers predicting non-response for patients receiving either Mepolizumab or Omalizumab are dominated by genes expressed in pDCs. (A) Venn diagram analysis for common biomarkers among 4 different comparisons of the cell type-gene features that have univariate ROC curve AUCs ≥ 0.8 for predicting non-response in patients receiving either Mepolizumab or Omalizumab (n=31). Biomarker sensitivity and specificity analysis by ROC curve for (B) pDCs-expressing *ANXA2* and (C) pDCs-expressing *DCAF5*, with 90% confidence intervals (blue shading). (D) Random forest analysis of top biomarkers using the mean decrease in the Gini coefficient as the metric of each features contribution to the prediction of non-response from the model with the top pDC gene markers in red box. (E) GO term analysis of the biological and molecular processes enriched in the genes that were most linked to non-response. (F) Volcano plot of fold change and p value for pDCs genes linked to non-response. Differential expression analysis as log2 fold change with p values indicated for the top and most consistent biomarkers predicting non-response (G) pDC-expressing *DCAF5*. ROC; receiver operating characteristic (ROC), AUC; area under the curve. Adj-CR; age/sex adjusted data for Cell Ranger, Non-adj-CR; non-adjusted data for Cell Ranger, Adj-AF; age/sex adjusted data for Alevin Fry, Non-adj-AF; non-adjusted data for Alevin Fry, GO; gene ontology.

### Individual clinical outcomes ACQ5, frequency of exacerbations, and OCS use are correlated with multiple lymphocyte subtype-gene combinations

We next examined how cell-gene combinations may correlate with individual clinical outcome measurements. Both a univariate and multivariate approach were taken, including using linear regression models and a random forest methodology to attribute the importance of individual cell-gene features within this model. Genes expressed in several types of lymphocytes ranged in their r^2^ linear correlation from 0.35 to 0.53 for the ACQ5 asthma symptom score, including genes in γδ T cells, MAIT cells, CD8+ T cells and B cells (Supplementary Figure 1A). Likewise, these cell-gene combinations were similarly the highest relative importance in a random forest model as measured by %IncMSE (Supplementary Figure 1B and Supplementary Table 2). The cell-gene combination with the strongest correlation with ACQ5 was γδ T cells-expressing *NUSAP1* (r2 = 0.53, p<0.000001). The higher the expression of this genes in these cells the greater the improvement in asthma symptoms. This suggests that about 53% of the variability in ACQ5 outcomes for patients receiving either Mepolizumab or Omalizumab could be explained by γδ T cell expression of *NUSAP1*. The majority of genes showing strongest linear correlations with the individual outcomes of frequency of exacerbation (Supplementary Figure 1C-D) and OCS use (Supplementary Figure 1E-F) over 6 months were expressed in pDCs, as well as genes in CD34+ progenitors, γδ T cells, B cells and NK cells (r^2^ ranging from 0.21-0.46 for the cell gene-combination displayed in Supplementary Figure 1C and E). Similar findings were present in the random forest model where the %IncMSE showed the highest relative importance in these models were an array of genes expressed in pDCs. (Supplementary Figure 1D and F and Supplementary Table 2). Overwhelmingly it is genes expressed in some of the less common lymphocyte subtypes that have the strongest linear correlations with the individual clinical outcomes ACQ5, OCS use and asthma exacerbations in 6 months. Two of these cell types (MAIT cells and CD34+ progenitors; Figure 6) were also the strongest predictors of the categorical outcome, clinical remission, which acts as the overall cumulative measure of change in ACQ5, OCS use, and exacerbations.

Use of a multivariate model builds on these top univariate predictions by computing on increasing numbers of features (k = 10, 25, 50, 100) selected from the top 500 ranked list of univariate cell-gene combinations based on differential expression FDRs. A 3:1 ratio of cases was reserved for training/testing. Regression is LASSO penalised, to observe consistent results. For ACQ5 this data reveals that near optimal performance was reached for most metrics (R2, RMSE; root mean squared error, and Huber_loss) with 25 cell-gene features using either a linear regression (LR) or random forest (RF) model (Supplementary Figure 2A). This produced an r^2^ of 0.97 using these 25 features explaining close to 100% of the variability in ACQ5 patient outcomes for Mepolizumab or Omalizumab (Supplementary Figure 2B). The multivariate random forest model (1000 trees) also showed strong performance with 25 cell type-gene features (listed) explaining the ACQ5 patient to patient outcomes with the decrease in Gini coefficient being used to determine relative importance of individual features within the multivariate model (Supplementary Figure 2C and Supplementary Table 2). Finally, SHapley Additive exPlanations, SHAP, was used to explain the output of the multivariate linear regression and random forest models (Supplementary Figure 2D-F and Supplementary Table 2). This approach is an alternative method of attributing importance to different components of the 25cell type-gene features and their relative weight in contribution to the overall performance of the multivariate model explaining variation in ACQ5. In this manner if a multi-component biomarker panel was to be designed these would be the features that contribute the most importance to the predictions of patient ACQ5 outcome.

## Discussion

Severe asthma with persistent type 2 inflammation, despite optimal treatment with inhaled corticosteroids, is a key disease endotype, which will benefit from treatment with biologic therapy. Within this response though there is substantial heterogeneity, with 20-30% of patients having a response of such significant magnitude that they can be considered to be so well controlled that clinicians are now proposing that therapy can induce clinical disease remission, though for the majority the response is not as marked, while 10-20% fail to respond and require switching to another biologic agent [11, 12, 27]. To better improve outcomes what is now required is precision use of these biologic therapies through biomarkers that can predict both ends of the spectrum, that is, non-response and clinical remission. The ability to predict the latter could be used in clinical decision-making to identify patients with the greatest clinical benefit and potentially to expand access to patients earlier in their disease process, prior to their exposure to significant morbidity, the long term consequences of OCS, and the associated health care costs. The ability to predict clinical remission would change the cost-benefit paradigm for severe asthma and biologic therapy considerably.

In order to address these questions we conducted a biomarker discovery study using blood from patients with severe type 2 asthma, prior to therapy initiation to be linked to their clinical outcomes after 6 months of either Meplolizumab or Omalizumab treatment. Our study utilised multiple bioinformatic analysis approaches with scRNAseq technology, which has a much higher resolution than traditional transcriptomic sequencing approaches to identify conserved biomarkers. scRNAseq can identify gene expression on a per cell basis in blood cells and so eliminates the major problems of using blood samples from a relatively homogenous population (severe asthma patients that meet strict eligibility criteria) where bulk transcriptomics causes an averaging effect of the gene expression among diverse cell types. scRNAseq can thus be used to identify blood cell biomarkers that can then be validated at the protein and cell type-specific level using antibody assays through flow cytometry or blood smear and immunofluorescence. Our study consistently identified ∼20 different types of WBCs across patients using two different bioinformatic approaches with CellRanger and AlevinFry. These methods reliably detected across patients the most abundant cell types, neutrophils and monocytes, as well as the rare cell types such as CD34+ multipotent progenitors, and basophils. The latter is another feature that is absent with traditional omic approaches. As might be expected in a relatively homogenous population of samples, the total cell type numbers in blood taken prior to therapy were not significantly different based on treatment outcome. We found that none of the baseline characteristics or cell or molecular markers currently used to assess asthma (FeNO, blood eosinophils, IgE) were accurate predictors of clinical remission or non-response (all below 0.76 ROC AUC).

Analysing gene expression on a per cell basis in the blood creates a higher resolution to identify biomarkers. This approach enabled more than 100 cell type-gene features to be identified with strong performance on ROC sensitivity and specificity analysis with AUC > 0.8. These features were identified across analysis pipeline methodologies and so we then determined cell type-gene features that were common to all analyses and therefore more likely reproducible biomarkers. The outcome of this was the delineation of *RPS26* (a ribosomal protein indicator of cellular activity) expressed in both CD34+ progenitors and circulating MAIT cells as excellent individual predictors of clinical remission (ROC AUC ∼0.9). These data are intriguing given these immune cell types are not among the most commonly associated with asthma phenotypes or pathogenesis, nor are they immediately apparent to be linked to allergic IgE or IL-5 pathway biology. However, there is evidence in the literature from preclinical models as well as patient samples for a role for these two cell types in asthma [28–31]. Given the results of this study demonstrating the strength of their association with clinical remission in severe asthma biologic therapy, there is a need for greater understanding of whether they are playing a functional role in clinical outcomes and what such a mechanism might involve.

This study is one of the first investigations using any unbiased global omics approach to identify biomarkers that predict clinical remission for severe asthma patients receiving biologic therapies. Furthermore, to our knowledge, this is the first study to utilise scRNAseq to investigate biomarkers in severe asthma treatment response, and the first study to identify a conserved single biomarker that accurately predicts outcome for multiple different biologic therapies (Mepolizumab and Omalizumab). We next investigated features linked to non-response to either therapy and discovered strong involvement of pDCs genes. These genes were predominately down-regulated in the blood of patients prior to therapy initiation who later went on to be non-responders. This pDC signature had multiple gene features with a 100% accuracy prediction. Overall, this generally lower pDC gene activity in non-responders appears to fit with the fact that this is a key anti-viral cell type known to play an important role in asthma exacerbations [32–34] and therefore lower activity in pDCs could be a likely cause of more exacerbations, leading to more OCS in severe asthma patients. Furthermore, pDCs have also been shown to play a role in modulating the IL-5 and IgE pathways involved in Omalizumab and Mepolizumab mechanisms of action [35, 36]. Similarly, the strongest correlation with individual clinical outcomes such as changes in OCS use, and exacerbations were also dominated by pDC genes likely based on this same biology of this interferon-producing cell type and their key role in antiviral responses. Virus infections remain the most common trigger of acute asthma, intriguingly treatment of exacerbation prone children with asthma with omalizumab not only reduced exacerbation risk, but also improved peripheral blood monocyte interferon responses to rhinovirus [37].

This is the first study that identifies novel cell type-gene biomarkers with high predictive accuracy (90-100%) to predict clinical remission to biologic therapy in people with severe type 2 asthma. This work justifies the design of specific protein-antibody assays (flow cytometry or blood smear immunofluorescence) to measure the protein level of these gene features in these specific cell types (e.g. CD34+ progenitors, MAIT cells, pDCs) that could act as more practical prognostic biomarker tests. This would then enable more efficient and cost-effective future validation of these biomarkers in larger cohort studies. The ability to predict clinical remission and other clinical outcomes for biologic therapies holds great promise for more effective precision medicine in severe asthma.

## Data Availability

Select de-identified data produced in the present study are available upon reasonable request to the authors

