## Supplementary materials for "Single cell RNA-seq discovery of blood biomarkers predicting treatment outcome in severe asthma patients"

### Supplementary Material

#### Supplementary Table 1

| Clinical Remission | ROC AUC | p value |
|---|---|---|
| FeNO baseline | 0.63 | 0.26 |
| Blood eosinophils baseline | 0.61 | 0.36 |
| Serum IgE baseline | 0.53 | 0.81 |
| % predicted FEV1 baseline | 0.59 | 0.44 |
| % predicted FVC baseline | 0.72 | 0.07 |
| OCS use baseline | 0.67 | 0.18 |
| ACQ5 baseline | 0.67 | 0.14 |
| Exacerbations baseline | 0.62 | 0.29 |
| Age | 0.71 | 0.07 |
| **Non-response Vs Response** | **ROC AUC** | **p value** |
| FeNO baseline | 0.74 | 0.12 |
| Blood eosinophils baseline | 0.58 | 0.55 |
| Serum IgE baseline | 0.76 | 0.07 |
| % predicted FEV1 baseline | 0.56 | 0.71 |
| % predicted FVC baseline | 0.62 | 0.58 |
| OCS use baseline | 0.7 | 0.2 |
| ACQ5 baseline | 0.67 | 0.25 |
| Exacerbations baseline | 0.63 | 0.43 |
| Age | 0.6 | 0.47 |
| **Change in ACQ5 Outcome** | **r2** | **p value** |
| FeNO baseline | 0.01 | 0.61 |
| Blood eosinophils baseline | 0.11 | 0.55 |
| Serum IgE baseline | 0.005 | 0.7 |
| % predicted FEV1 baseline | 0.02 | 0.48 |
| % predicted FVC baseline | 0.003 | 0.77 |

| | | |
|---|---|---|
| OCS use baseline | 0.008 | 0.65 |
| ACQ5 baseline | **0.4** | **0.0001** |
| Exacerbations baseline | 0.02 | 0.41 |
| Age | 0.01 | 0.69 |
| **Change in OCS Outcome** | **r2** | **p value** |
| FeNO baseline | 0.12 | 0.09 |
| Blood eosinophils baseline | 0 | 0.97 |
| Serum IgE baseline | 0.012 | 0.6 |
| % predicted FEV1 baseline | 0.07 | 0.2 |
| % predicted FVC baseline | 0.12 | 0.11 |
| OCS use baseline | **0.97** | **0.0001** |
| ACQ5 baseline | 0.04 | 0.33 |
| Exacerbations baseline | **0.48** | **0.0001** |
| Age | 0.009 | 0.65 |
| **Change in % predicted FEV1 Outcome** | **r2** | **p value** |
| FeNO baseline | 0.01 | 0.58 |
| Blood eosinophils baseline | 0.004 | 0.76 |
| Serum IgE baseline | 0.01 | 0.58 |
| % predicted FEV1 baseline | 0.16 | 0.08 |
| % predicted FVC baseline | 0.13 | 0.07 |
| OCS use baseline | 0.07 | 0.2 |
| ACQ5 baseline | 0.06 | 0.22 |
| Exacerbations baseline | 0.004 | 0.74 |
| Age | 0.11 | 0.08 |
| **Change in Exacerbations Outcome** | **ROC AUC** | **p value** |
| FeNO baseline | 0.13 | 0.06 |
| Blood eosinophils baseline | 0.01 | 0.58 |

| | | |
|---|---|---|
| Serum IgE baseline | 0.02 | 0.48 |
| % predicted FEV1 baseline | 0.06 | 0.22 |
| % predicted FVC baseline | 0.12 | 0.09 |
| OCS use baseline | **0.47** | **0.0001** |
| ACQ5 baseline | 0.05 | 0.25 |
| Exacerbations baseline | **0.96** | **0.0001** |
| Age | 0.04 | 0.28 |
| **% change in ACQ5 Outcome over baseline** | **r2** | **p value** |
| FeNO baseline | 0.002 | 0.83 |
| Blood eosinophils baseline | 0.0006 | 0.9 |
| Serum IgE baseline | 0.00009 | 0.96 |
| % predicted FEV1 baseline | 0.02 | 0.43 |
| % predicted FVC baseline | 0.13 | 0.06 |
| OCS use baseline | 0.001 | 0.87 |
| ACQ5 baseline | 0.1 | 0.09 |
| Exacerbations baseline | 0.01 | 0.56 |
| Age | 0.01 | 0.6 |
| **% change in OCS Outcome over baseline** | **r2** | **p value** |
| FeNO baseline | 0.0003 | 0.93 |
| Blood eosinophils baseline | 0.04 | 0.3 |
| Serum IgE baseline | 0.02 | 0.4 |
| % predicted FEV1 baseline | 0.07 | 0.2 |
| % predicted FVC baseline | 0.006 | 0.72 |
| OCS use baseline | 0.02 | 0.48 |
| ACQ5 baseline | 0.008 | 0.66 |
| Exacerbations baseline | 0.03 | 0.43 |
| Age | 0.02 | 0.55 |

| % change in % of predicted FEV1 Outcome over baseline | r2 | p value |
|---|---|---|
| FeNO baseline | 0.02 | 0.5 |
| Blood eosinophils baseline | 0.0002 | 0.94 |
| Serum IgE baseline | 0.004 | 0.75 |
| % predicted FEV1 baseline | **0.28** | **0.004** |
| % predicted FVC baseline | **0.26** | **0.008** |
| OCS use baseline | 0.13 | 0.09 |
| ACQ5 baseline | **0.16** | **0.03** |
| Exacerbations baseline | 0.05 | 0.25 |
| Age | 0.05 | 0.25 |
| **% change in Exacerbations Outcome over baseline** | **ROC AUC** | **p value** |
| FeNO baseline | 0.02 | 0.44 |
| Blood eosinophils baseline | 0.001 | 0.85 |
| Serum IgE baseline | 0.002 | 0.8 |
| % predicted FEV1 baseline | 0.01 | 0.58 |
| % predicted FVC baseline | 0.02 | 0.46 |
| OCS use baseline | 0.03 | 0.4 |
| ACQ5 baseline | 0.02 | 0.44 |
| Exacerbations baseline | 0.12 | 0.07 |
| Age | 0.001 | 0.87 |

**Supplementary Table 2: cell type identifiers for random forest plots**

| | |
|---|---|
| CL:0000043: | mature basophil |
| CL:0000049: | common myeloid progenitor |
| CL:0000050: | megakaryocyte-erythroid progenitor cell |
| CL:0000233: | platelet |

| | |
|---|---|
| CL:0000236: | Other B cell |
| CL:0000787: | memory B cell |
| CL:0000788: | naive B cell |
| CL:0000451: | dendritic cell |
| CL:0000576: | monocyte |
| CL:0000623: | natural killer cell |
| CL:0000624: | CD4+ alpha-beta T cell |
| CL:0000625: | CD8+ alpha-beta T cell |
| CL:0000771: | eosinophil |
| CL:0000775: | neutrophil |
| CL:0000784: | plasmacytoid dendritic cell |
| CL:0000798: | gamma-delta T cell |
| CL:0000860: | classical monocyte |
| CL:0000875: | non-classical monocyte |
| CL:0000940: | mucosal invariant T cell |
| CL:0002043: | CD34+ multipotent progenitor cell |

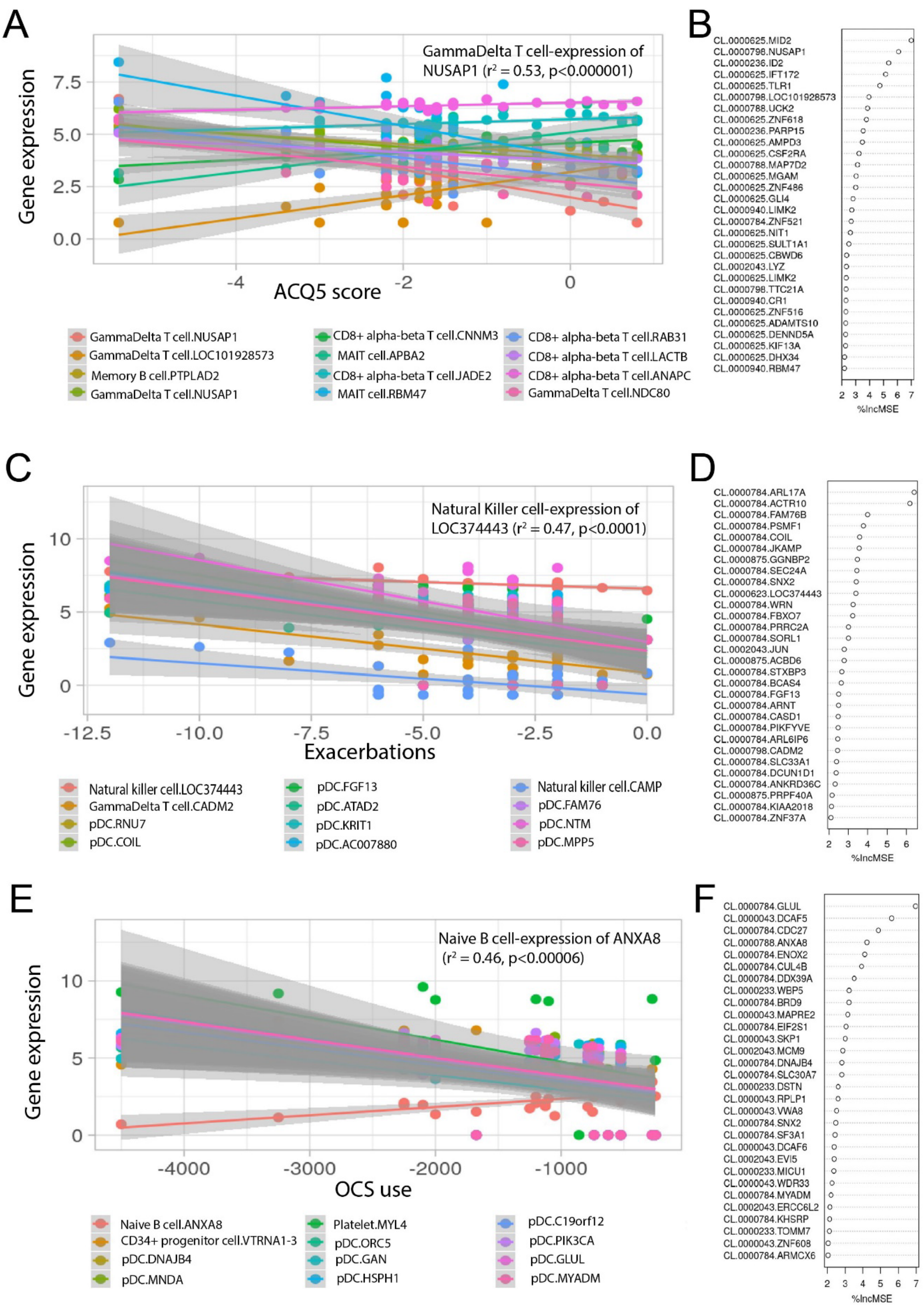
A
GammaDelta T cell-expression of NUSAP1 (r² = 0.53, p<0.000001)
Gene expression
ACQ5 score
GammaDelta T cell.NUSAP1
GammaDelta T cell.LOC101928573
Memory B cell.PTPLAD2
GammaDelta T cell.NUSAP1
CD8+ alpha-beta T cell.CNNM3
MAIT cell.APBA2
CD8+ alpha-beta T cell.JADE2
MAIT cell.RBM47
CD8+ alpha-beta T cell.RAB31
CD8+ alpha-beta T cell.LACTB
CD8+ alpha-beta T cell.ANAPC
GammaDelta T cell.NDC80
B
CL.0000625.MID2
CL.0000796.NUSAP1
CL.0000236.ID2
CL.0000625.IFT172
CL.0000625.TLR1
CL.0000798.LOC101928573
CL.0000788.UCK2
CL.0000625.ZNF618
CL.0000236.PARP15
CL.0000625.AMPD3
CL.0000625.CSF2RA
CL.0000788.MAP7D2
CL.0000625.MGAM
CL.0000625.ZNF486
CL.0000625.GLI4
CL.0000940.LIMK2
CL.0000784.ZNF521
CL.0000625.NIT1
CL.0000625.SULT1A1
CL.0000625.CBWD6
CL.0002043.LYZ
CL.0000625.LIMK2
CL.0000798.TTC21A
CL.0000940.CR1
CL.0000625.ZNF516
CL.0000625.ADAMTS10
CL.0000625.DENND5A
CL.0000625.KIF13A
CL.0000625.DHX34
CL.0000940.RBM47
%IncMSE
C
Natural Killer cell-expression of LOC374443 (r² = 0.47, p<0.0001)
Gene expression
Exacerbations
Natural killer cell.LOC374443
GammaDelta T cell.CADM2
pDC.RNU7
pDC.COIL
pDC.FGF13
pDC.ATAD2
pDC.KRIT1
pDC.AC007880
Natural killer cell.CAMP
pDC.FAM76
pDC.NTM
pDC.MPP5
D
CL.0000784.ARL17A
CL.0000784.ACTR10
CL.0000784.FAM76B
CL.0000784.PSMF1
CL.0000784.COIL
CL.0000784.JKAMP
CL.0000875.GGNBP2
CL.0000784.SEC24A
CL.0000784.SNX2
CL.0000623.LOC374443
CL.0000784.WRN
CL.0000784.FBXO7
CL.0000784.PRRC2A
CL.0000784.SORL1
CL.0002043.JUN
CL.0000875.ACBD6
CL.0000784.STXBP3
CL.0000784.BCAS4
CL.0000784.FGF13
CL.0000784.ARNT
CL.0000784.CASD1
CL.0000784.PIKFYVE
CL.0000784.ARL6IP6
CL.0000798.CADM2
CL.0000784.SLC33A1
CL.0000784.DCUN1D1
CL.0000784.ANKRD36C
CL.0000875.PRPF40A
CL.0000784.KIAA2018
CL.0000784.ZNF37A
%IncMSE
E
Naive B cell-expression of ANXA8 (r² = 0.46, p<0.00006)
Gene expression
OCS use
Naive B cell.ANXA8
CD34+ progenitor cell.VTRNA1-3
pDC.DNAJB4
pDC.MNDA
Platelet.MYL4
pDC.ORC5
pDC.GAN
pDC.HSPH1
pDC.C19orf12
pDC.PIK3CA
pDC.GLUL
pDC.MYADM
F
CL.0000784.GLUL
CL.0000043.DCAF5
CL.0000784.CDC27
CL.0000788.ANXA8
CL.0000784.ENOX2
CL.0000784.CUL4B
CL.0000784.DDX39A
CL.0000233.WBP5
CL.0000784.BRD9
CL.0000043.MAPRE2
CL.0000784.EIF2S1
CL.0000043.SKP1
CL.0002043.MCM9
CL.0000784.DNAJB4
CL.0000784.SLC30A7
CL.0000233.DSTN
CL.0000043.RPLP1
CL.0000043.VWA8
CL.0000784.SNX2
CL.0000784.SF3A1
CL.0000043.DCAF6
CL.0002043.EVI5
CL.0000233.MICU1
CL.0000043.WDR33
CL.0000784.MYADM
CL.0002043.ERCC6L2
CL.0000784.KHSRP
CL.0000233.TOMM7
CL.0000043.ZNF608
CL.0000784.ARMCX6
%IncMSE

**Supplementary Figure 1. Cell type-gene feature correlations with individual clinical outcomes ACQ5, OCS use, and frequency of exacerbations following either Mepolizumab or Omalizumab treatment.** (A-C) Univariate linear regression analysis with top 12 cell type gene features shown with importance measured by $r^2$ values, and (B-F) univariate random forest model with top 30 cell-type gene feature importance measured by mean decrease accuracy (% IncMSE) for individual clinical outcomes change in ACQ5 (A-B), change in frequency of exacerbations (C-D), and change in OCS use (E-F). Outcomes are those combined for patients receiving either Mepolizumab or Omalizumab (n=31). Top feature for each clinical outcome in (A-C) indicated with $r^2$ and p value in the text box.

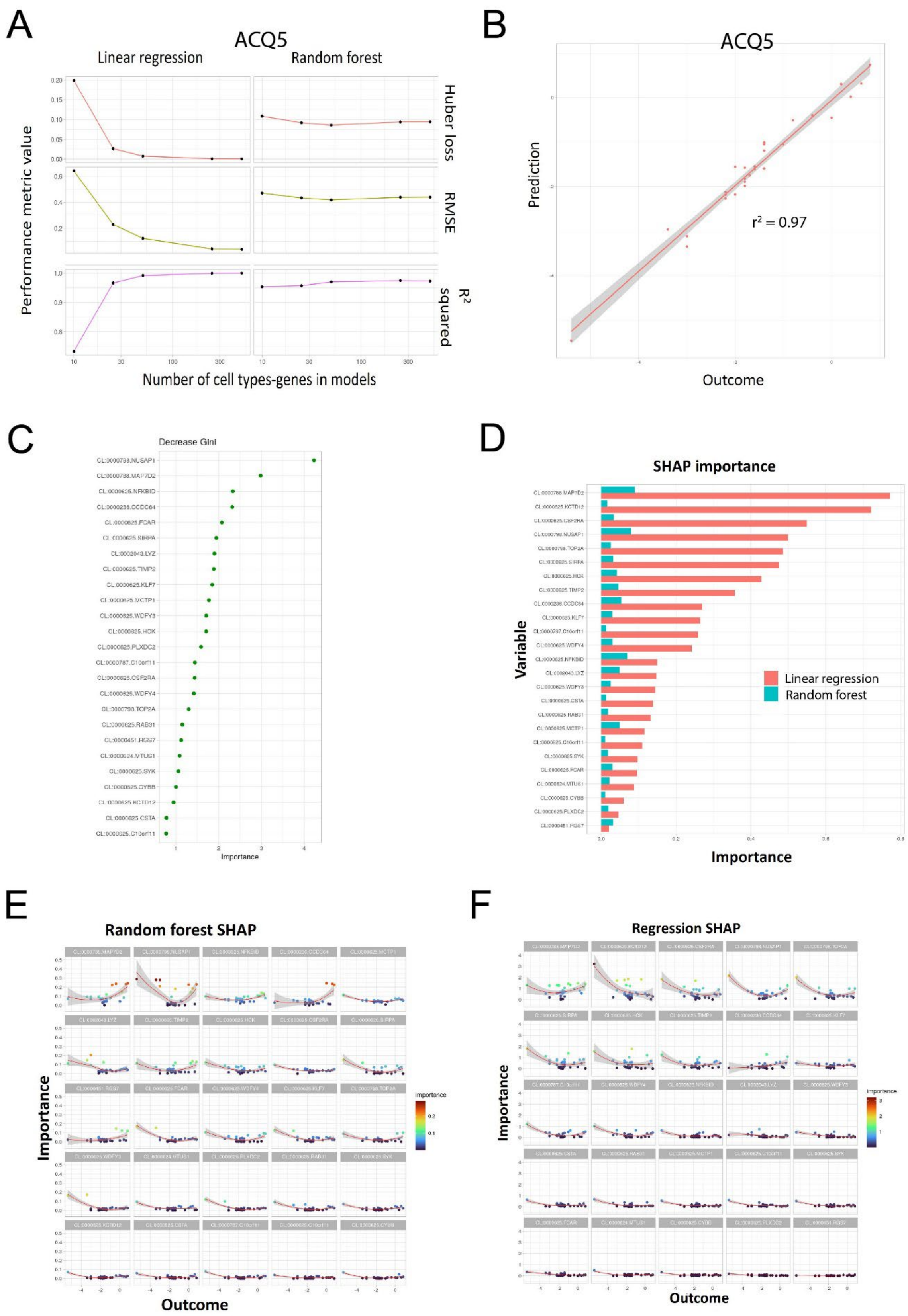
A
ACQ5
Linear regression
Random forest
Huber loss
RMSE
R2 squared
Performance metric value
Number of cell types-genes in models
B
ACQ5
Prediction
Outcome
r2 = 0.97
C
Decrease Gini
Importance
D
SHAP importance
Variable
Importance
Linear regression
Random forest
E
Random forest SHAP
Importance
Outcome
F
Regression SHAP
Importance
Outcome

**Supplementary Figure 2. Multivariate model analysis of cell type-gene features that correlate with asthma symptoms by ACQ5 following treatment with either Mepolizumab or Omalizumab.** (A) Linear regression and random forest models display increasing model performance (r2, RMSE, and Huber_loss) with increased number of cell type-gene features. (B) overall linear regression of 25 cell-type gene features with change in ACQ5 before and after 6 months treatment. (C) Random forest model with each of the 25 cell-type gene features relative importance indicated by decrease in the Gini coefficient. (D-F) SHAP feature analysis explaining the importance of each of the 25 cell type-gene features in the regression and random forest models. RMSE; root mean square error, SHAP; SHapley Additive exPlanations.
